# Impending Heart Failure : An Artificial Intellectual Reality

**DOI:** 10.1101/2024.08.12.24311907

**Authors:** Abhijit Ray

**Affiliations:** Chief Scientific Officer, HAIF Inc.

## Abstract

**Background:** Having a prevalence of almost 64 million patients globally, Heart Failure (HF) remains a leading cause of cardiac death with high morbidity and mortality rates despite constant updates in diagnostic and therapeutic measures. One of the main reasons behind this might be a delay in initiating treatment.

**Purpose:** To obtain a method of screening patients for Heart Failure even before they develop symptoms.

**Methods:** An Artificial Intelligence-based algorithm, named Heart Failure Predictor (HFP), born from mathematical calculations of a patented formula, came up with a cutting-edge solution that can predict the chances of HF in patients not suffering from any symptoms of Heart Failure. HFP was applied to data from the Framingham Heart Study as a retrospective analysis. LVEF and NT pro-BNP levels were used as a method of correlation. Asymptomatic patients who had been followed up extensively for at-least 24 months were included, while patients already diagnosed with HF were excluded.

**Results:** Data from 20896 patients were analysed.

17 out of 1230 were false positive while 31 out of 19660 were false negative.

Thus, HFP has a Positive Predictive Value of 98.6% and a Specificity of 99.9%.

**Discussion:** Early screening and detection may lead to vast improvements in HF treatment efficacy. HFP fills this need with a new classification of HF, “Impending Heart Failure”, and synonymously “Ray’s Disease”.

Apart from screening HF patients, HFP can also be used for continuous monitoring of LVEF in cardiac beds, bringing about a revolution in the cardiac space.

## Background

Heart Failure has been raging across the globe as a cardiac pandemic and is a staggering clinical and public health problem, associated with significant mortality and morbidity; not to mention healthcare-related expenditures, particularly amongst those aged 65 and above.

Heart Failure (HF) has been defined as a clinical syndrome with symptoms and/or signs caused by a structural and/or functional cardiac abnormality and corroborated by elevated natriuretic peptide levels and/or objective evidence of pulmonary or systemic congestion. A new and revised classification of HF according to left ventricular ejection fraction (LVEF) includes HF with reduced EF (HFrEF): HF with an LVEF of ≤40%; HF with mildly reduced EF (HFmrEF): HF with an LVEF of 41% to 49%; HF with preserved EF (HFpEF): HF with an LVEF of ≥50%; and HF with improved EF (HFimpEF) : HF with a baseline LVEF of ≤40%, a ≥10-point increase from baseline LVEF, and a second measurement of LVEF of >40%^1^.

Thus, LVEF is clearly the single-most important parameter for diagnosis and prognosis of Heart Failure cases. According to the American Heart Association, LVEF is measured (expressed as a percentage) of how much blood the left ventricle pumps out with each contraction^2^. It is the most common method for assessment of systolic function. In many cardiovascular emergencies including Acute Myocardial Infarction, Complete Heart Block and Acute Congestive Cardiac Failure, there is often an urgency to measure LVEF since treatment depends on it.

At present, the standard method of assessing LVEF is by conducting an Echocardiography, using the modified Quinones Equation^3^. Hence, LVEF can only be assessed as snapshots rather than as a continuous parameter. LVEF can be determined using several invasive and non-invasive imaging modalities, either subjectively by visual estimation or objectively by quantitative methods i.e. echocardiography, computed tomography (CT), magnetic resonance imaging (MRI), gated myocardial perfusion imaging with either single-photon emission computed tomography (SPECT) or positron emission tomography (PET) and gated equilibrium radionuclide angiography (commonly referred to as multiple-gated acquisition [MUGA] scan)^4^. LVEF can also be measured non-invasively using the first-pass radionuclide technique, but this technique is rarely performed in the current era. However, these techniques do not possess the advantage of continuous monitoring. Apart from chronic factors like Hypertension, Diabetes, smoking etc, multiple acute factors also affect LVEF including

- Sympathetic stimulation
- Blood volume
- Respiration
- Environmental temperature^5^

Due to these, LVEF is a dynamic entity whose continuous monitoring might be extremely beneficial for treatment instead of snapshot values of LVEF (as is available from current LVEF measurement techniques).

Other problems faced by the current methods of LVEF measurement (specially Echocardiography) are :

1. LVEF estimation from Echocardiography is subject to observer bias
2. Requiring specialised equipment (Echocardiography machine) and trained personnel for obtaining the results
3. It is costly, specially in cases where multiple readings are required Because of all these factors, a great percentage of the Heart Failure population remains undiagnosed.

This paper aims to present a predictive screening mechanism for Heart Failure which is quick, accessible and cheap.

## Methodology

A patent issued by the United States Patent and Trademark Office (USPTO)^6^ provides a mathematical formula that calculates LVEF from an ECG recording. A software named Heart Failure Predictor (HFP) took this basic formula and applied AI/ML technology to come up with a cutting edge-solution that can predict the chances of Heart Failure in patients not suffering from any symptoms of Heart Failure.

HFP naturally works on digital ECG data. Instead of accessing digital data directly from the ECG machines, ECG paper reports were scanned and then converted from analogue to digital data^7^. The algorithm works on linear regression model^8^ using inputs from not only ECG recordings, but also other factors on which Heart Failure is diagnosed including Blood Pressure measurements, Heart Rate, SpO_2_ LVEF and NT pro-BNP values, chamber sizes, and pulmonary arterial systolic pressure (PASP)^9^. Retrospective analyses were done amongst known Heart Failure patients to evaluate and input all these factors into the machine learning program and create a reverse timeline until the origin of the disease in that patient. After doing the same for 150 data points, the machine could develop its own predictive algorithm which has been termed Heart Failure Predictor.

For proof of concept, 136 patients were taken as a prospective trial and followed up at regular intervals and showed the algorithm had over 95% success rate at predicting heart failure cases. Then, for a retrospective analysis, data from the Framingham Heart Study was requested and analysed.

### Inclusion criteria

Adults who had been closely monitored with ECG, Echocardiography and NT pro-BNP for at least 24 months.

### Exclusion criteria

Patients already suffering from various stages of Heart Failure were excluded from this analysis.

Using these criteria, a total of 20896 patients’ data were analysed. Each analysis contained 3 steps:

1. Measuring LVEF from the ECGs of the patients
2. Using a regression machine learning model to predict risk of Heart Failure
3. Corroborating the findings with future data (LVEF measurements and NT pro-BNP levels) of the same patient

## Results

Of the 20896 patients, the algorithm predicted 1236 patients to be at “High Risk” of heart failure. Correlating the follow-up reports of these patients revealed 1219 of them developed heart failure at some point within the next 24 months. 1206 of these patients had reduction in LVEF coupled with manifold increase in NT pro-BNP levels and were thus diagnosed as HFrEF. 13 out of the 1219 patients had maintained LVEF values, however were diagnosed as HFpEF due to increased NT pro-BNP levels adding to other symptoms.

The remaining 19660 patients were labelled as “no immediate risk” by the algorithm, wherein 31 patients developed Heart Failure in the following 24 months while the other 19629 remained unaffected.

**Table 1.** Tabulation of Results.

|  | Developed HFrEF | Developed HFpEF | No Heart Failure |
| --- | --- | --- | --- |
| HFP High Risk | 1206 | 13 | 17 |
| HFP Low Risk | 8 | 23 | 19629 |

*Table 1 : Tabulation of Results*
|  | Developed HFrEF | Developed HFpEF | No Heart Failure |
| --- | --- | --- | --- |
| HFP High Risk | 1206 | 13 | 17 |
| HFP Low Risk | 8 | 23 | 19629 |

These results show that the HFP algorithm has, for Heart Failure as a whole :

I. Specificity of 99.9%
II. Sensitivity of 97.5%
III. Positive Predictive Value of 98.6%
IV. Negative Predictive Value of 99.8%

## Discussion

With the rate of advancement of modern software technology, the merge of software with healthcare has been inevitable. Softwares associated with digital radiology, machines for faster processing of lab samples, inclusion of various softwares including ECG and sound recording mechanisms inside stethoscopes; these are just a few examples of how modern healthcare has been advancing rapidly with the help of softwares and algorithms.

This study shows that with the use of modern-day technology, Heart Failure can be predicted even before the symptoms start developing. Since it is an irreversible disease^10^, early detection or screening can be extremely beneficial to improve morbidity and mortality rates, not to mention the healthcare-associated expenses.

HFP fills in this need with a new classification of Heart Failure which we term as “Impending Heart Failure”, and synonymously “Ray’s Disease”.

The American Heart Association has staged Heart Failure into 4 classes (A,B,C and D) where Stage B (Pre-Heart Failure)^11^ is defined as patients without any symptoms of Heart Failure but have risk factors like structural heart disease, increased filling pressures in the heart, etc.

Why the need for this new stage of Impending Heart Failure while Pre-Heart Failure patients have already been defined and categorised? Mostly for two reasons

I. <u>Definite progression</u> : while Pre-Heart Failure patients are at much higher risk of developing Heart Failure than the general population, they may or may not progress to Stage C (Symptomatic Heart Failure). However, HFP having a positive predictive value of 98.6% translates to Impending Heart Failure patients progressing to Symptomatic Heart Failure in at least 98.6% cases.
II. <u>Timeline</u> : while in today’s world it is very difficult to provide a patient quantitative report on when he or she shall go into Symptomatic Heart Failure, HFP provides a certain timeline which both patients and clinicians can rely upon to start as rigorous management of the disease and/or the causes as they deem necessary.

## Conclusion

Apart from HFP’s usage as a Heart Failure screening and prediction, the algorithm can also be used for continuous LVEF monitoring in patients admitted with cardiac monitors (intensive care, emergency rooms etc). This real-time continuous monitoring may be extremely beneficial to manage not only HF related morbidity and mortality rates^12^, but also other diseases which affect haemodynamic stability. While the burden of Heart Failure is too big to bear for most countries across the world, very few institutions adapt new technology to improve patient outcomes^13^. How soon HFP will be adopted across the globe will be yet to be seen, but once it is, we are very optimistic about its results on the cardiology world at large.

## Data Availability

Data received from Framingham Heart Study.

